# Prognostic significance of circRNAs expression in breast carcinoma patients: A meta-analysis

**DOI:** 10.1101/2021.01.02.21249132

**Authors:** Zizhen Zhou, Xiancai Li, Dewu Liu

**Author notes:** **Correspondence to**: Dewu Liu, Department of Burns, The First Affiliated Hospital of Nanchang University. No.17, Yongwaizhengjie Road, Donghu District Nanchang 330006, Jiangxi, China; E-mail addresses. ZZZ and XCL contributed equally to this study. **Credit author statement** XC.L study concept and design, data collection; ZZ.Z data analyzing and article writing; DW.L Obtained funding and critically revised the manuscript. All of the authors reviewed the manuscript and approved the final version. **Declaration of Competing Interest** The authors declare that there are no conflicts of interest.

## Abstract

**Objective:** The aim of our study was to systematically evaluate the prognostic effects of various circrnas and to explore the prognostic value of circRNAs in breast cancer patients.

**Methods:** A systematical search was conducted on PubMed, Scopus, EMBASE, and the Cochrane Library databases. Eligible studies reporting on the association among circRNAs and prognostic values of breast cancer patients were included. Fixed-effects and random effects models were used to calculate the pooled hazard ratio values of overall survival and disease free survival. In addition, funnel plots were used to qualitatively analyze the publication bias.

**Results:** 28 studies were included in our meta-analysis. The pooled hazard ratio values of overall survival and disease free survival related to different circRNAs expression in breast cancer patients were 1.68 (1.44-1.97), 2.63 (1.95-3.53).We have identified a total of 28 circRNAs including 19 significantly up-regulated expression circRNAs and 9 significantly down-regulated expression circRNAs in BC(breast cancer) patients. Moreover, all of them revealed mechanisms and have the function of promoting or inhibiting the proliferation, metastasis or invasion of breast cancer cells by acting on the corresponding target.

**Conclusion:** Overall, specific circRNAs are significantly associated with the prognosis of BC patients and potentially eligible for the prediction of patients survival. It also provides a potential value for clinical decision-making development and may serve as a promising circRNAs-based target therapy waiting for further elucidation.

## 1. Introduction

Breast cancer (BC) is a common cancer that greatly affects globle women and is the leading cause of cancer death[1].The incidence of BC has been reported to be on the rise over the past decade, in addition to the fact that female patients continue to have the highest mortality[2]. It is also reported that there were 2,088,000 new cases of BC, and 626,000 women died of BC worldwide in 2018.Although the diagnosis and treatment strategies of BC have been improved and the overall survival time of BC patients is relatively long, the prognosis of BC patients cannot achieve the expected effect due to the complex pathogenesis of breast cancer, the molecular characteristics of BC and the diverse clinical manifestations of BC patients.Therefore, it is urgent to better understand the key signaling pathways and find new therapeutic targets for more effective treatment of BC in order to improve the prognosis of BC patients.

Circular RNAs (circRNAs), a novel class of ncRNAs with 3′- and 5′-ends covalently linked in a closed loop structure, are ubiquitous in all eukaryotic cells[3-5]. However, circRNAs discovered in the early stage has always been considered as not functional. With the development of RNA sequencing technology and bioinformatics, circRNA’s functionality has been gradually discovered and further studied[6]. Extensive studies[7, 8]have shown that circRNAs are abundant in eukaryotic cells and play a key role in human diseases, including BC, by binding to microrna (MIRS), interacting with RNA-binding proteins, and regulating transcription. CircRNAs can directly regulate protein expression through interaction with miRs, thereby promoting or inhibiting proliferation, invasion and metastasis of breast cancer cells [9]. Yang et al.[10]found that circ_0103552 expression significantly increased in breast cancer tissue samples and cells, and it was associated with clinical severity and poor prognosis, including advanced TNM stage and positive lymph node infiltration. It can directly play a carcinogenic role through the spongy action of Mir-1236. In addition, the upregulation of CIRc_0103552 can also promote cell growth, clonal formation, migration and invasion, and reduce apoptotic cells. A recent study [11]showed that the expression of circRNA_000554 in BC tissues was significantly lower than that in normal tissues, and the survival time of hsa_circRNA_000554 with low expression was shorter than that of circRNA_000554 with overexpression. In addition, they found that circRNA_000554 with overexpression inhibited EMT, cell invasion and migration during breast cancer by decreasing Mir-182 and increasing ZFP36.

To sum up, circRNAs expression may affect the prognosis of breast cancer.In our study, we performed a meta-analysis to summarize the prognostic significances of circRNAs in BC patients. Further prospective studies including more kinds of circRNAs are warranted in the future.

## 2. MATERIALS AND METHODS

### 2.1. Search strategy

A computerized literature search was performed in the PubMed, Scopus, EMBASE, and the Cochrane Library databases up to 25 november 2018. The search strategy of our study followed the terms such as: (a) “circRNA” or”circular RNA”; and (b) “breast cancer” or “breast carcinoma” or “breast tumor” or “breast carcinoma”.In addition, we hand□searched the references of all relevant articles one by one if it is necessary. When the important data were not available, we tried to contact researchers of certain articles.

### 2.2. Inclusion and exclusion criteria

The article selection used the following inclusion and exclusion criteria for each study. A study that is eligible for inclusion must meet the following criteria:(1) Case control study or cohort study, including case group and control group;(2) Breast cancer patients were diagnosed by pathologists;(3) Detection of circRNAs expression level, clinicopathological features, and prognosis of patients;(4)Overall survival (OS), disease-free survival (DFS), hazard ratio (HR), and 95 confidence interval (CI) can be obtained directly from the full text, or survival data can be extracted from kaplan-Meier survival curve.

Moreover, the exclusion articles all fitted the following criteria:(1)Studies not relevant to circRNA or BC;(2)Similar studies or repeated data in different articles;(3)Animal studies, comments, case reports, expert opinions, letters, etc.;(4)Without available data for analysis and the authors could not be contacted;(5) Non-English literature.

### 2.3. Data Extraction

The two authors (Xiancai Li and Zizhen Zhou) extracted data independently and got consensus finally. (A) baseline information for each study, including author, year of publication, nation of population enrolled, number of patients, circRNAs type,HR and 95% CI (OS/DFS), cut-off value, method, sample type, and follow-up. (B)we extracted the functions and targets related to the up-regulation and down-regulation of circRNA expression so as to illustrate the mechanism of circRNA’s effect on breast cancer prognosis.

### 2.4. Statistical analysis

statistical analysis was conducted by Stata version 12.0. hazard ratios (HRs) and 95% confidence intervals (CIs) were utilized to evaluate OS/DFS. The chi□square test and the I^2^ statistic were utilized to assess the between□study heterogeneity .P < 0. 01 was considered statistically significant. The heterogeneity among studies was calculated by Q and I^2^ tests. P > 0.10 in combination with I^2^ < 50% indicated low heterogeneity(note: I^2^ <25% suggests low heterogeneity; 25 ≦ I^2^ ≦ 75% suggests moderate heterogeneity; I^2^ >75% shows high heterogeneity),which should use fixed-effect models. Otherwise, random-effect models would be used finally. For some studies from which we could not extract HR and corresponding 95% CI (OS/DFS) directly, Engauge Digitizer 4.1 software was applied to obtain the necessary points and the relevant data from Kaplan-Meier survival curves, then HR and corresponding 95% CI were calculated by published methods proposed by Tierney et al[12]. Additionally, forest plots of the pooled HR values and funnel plots used to analyse qualitatively publication bias were presented.

## 3. Results

### 3.1. Search results

The study search is shown in the flow diagram (Figure 1). 592 relevant articles were collected during the database search. 194 duplicate literatures were excluded, and 370 literatures were excluded after reading the titles and full texts.To sum up, 28 eligible studies were included in the present study. All the selected studies included 27 for OS, 5 for DFS analysis.

**Figure 1:**
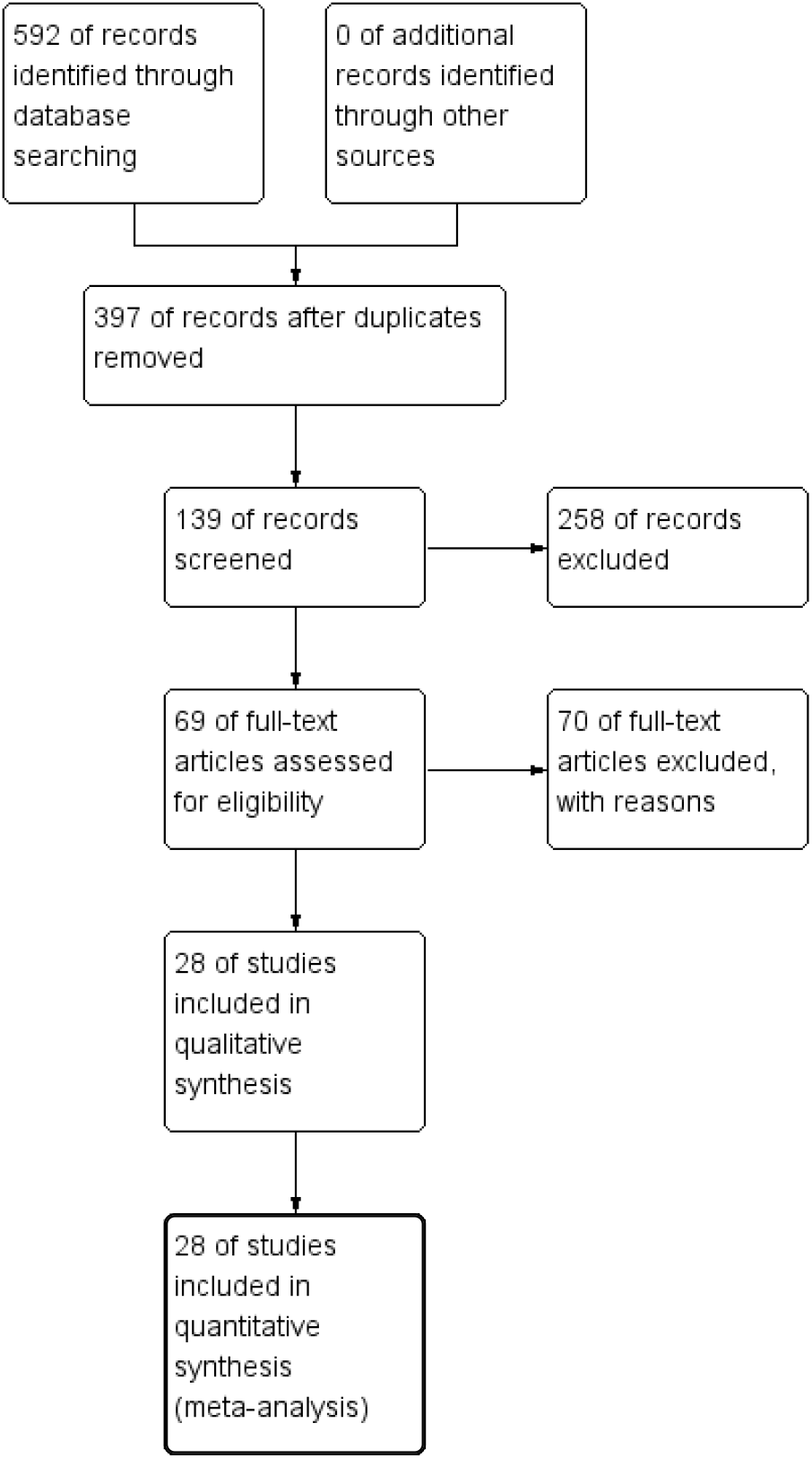
Study flow diagram.

### 3.2. Study characteristics

The main features of each eligible study are summarized in details (Tables 1 and 2). The years for publication ranged from 2018 to 2020. Most of the studies taken into account refer to Asian population, especially china.The range of sample size in each study was from 30 to 350. Cut-off values of high or low circRNA expression were mostly median or mean.The range of mean duration of follow□up was from 30 to 250 months.Moreover,Detection methods of circRNA expression were mainly quantitative real□time polymerase chain reaction (RT□PCR).Sample types were almost from tissues. As for clinical outcome indicators, 27 studies[10, 11, 13-34],[35-37]included overall survival (OS), 5 studies[19, 29, 31, 33, 38]included disease-free survival (DFS).As shown in Table 3, nineteen types of circRNAs were recognized as tumor promoters and nine were tumor suppressors. 28 circrnas have corresponding targets and can promote or inhibit the proliferation, invasion and metastasis of breast cancer cells.

**Table 1:** Characteristics of studies and different circRNAs expression related to OS in BC patients.

| study | CircRNA<br>( n=27 ) | year | Nation | Number<br>(n=3636) | OS |  | cut-off<br>value | Sample<br>types | Detecti<br>methoc |
| --- | --- | --- | --- | --- | --- | --- | --- | --- | --- |
|  |  |  |  |  | HR | 95%CI |  |  |  |
| Yang etal | circ_0103552↑ | 2019 | China | 57 | 2.32 | 1.03-5.23 | Mean | Tissue | RT-PC |
| Yan etal | hsa_circ_0072309 ↓ | 2019 | China | 32 | 1.73 | 0.36-8.42 | NR | Tissue | RT-PC |
| Xu etal | circ_0005230 ↑ | 2018 | China | 76 | 1.95 | 1.02-3.70 | Median | Tissue | RT-PC |
| Xu etal | Hsa_circ_001569 ↑ | 2019 | China | 145 | 3.03 | 1.33-4.24 | Median | Tissue | RT-PC |
| Wang etal | circUBE2D2 ↑ | 2019 | China | 80 | 1.39 | 0.12-16.01 | Median | Tissue | RT-PC |
| Wang etal | MYO9B ↑ | 2018 | China | 41 | 2.79 | 0.27-29.23 | NR | Tissue | RT-PC |
| Liang etal | circKDM4C ↓ | 2019 | China | 219 | 0.1687 | 0.0288-0.98 | 1.5FC | Tissue | RT-PC |
| Gao etal | hsa_circRNA_0006528<br>↑ | 2018 | China | 96 | 1.39 | 0.44-4.36 | Median | Tissue | RT-PC |
| Xie etal | hsa_circ_0004771 ↑ | 2019 | China | 51 | 1.49 | 0.83-2.68 | 1.0FC | Tissue | RT-PC |
| Zeng etal | ANKS1B ↑ | 2019 | China | 165 | 3.29 | 1.36-7.93 | Median | Tissue | RT-PC |
| Cao etal | circRNF20 ↑ | 2020 | China | 50 | 1.26 | 0.44-3.57 | NR | Tissue | RT-PC |
| Liang etal | BMPR2 ↓ | 2019 | China | 199 | 0.4539 | 0.275-0.746 | 2.0FC | Tissue | RT-PC |
| Liang etal | circCDYL ↑ | 2020 | China | 30 | 3.75 | 1.23-11.24 | Median | Tissue | RT-PC |
| Zhou etal | FBXL5 ↑ | 2019 | China | 150 | 1.73 | 0.41-7.33 | 2.0FC | Tissue | RT-PC |
| Song etal | CircHMCU ↑ | 2020 | China | 267 | 3.09 | 1.44-6.66 | Median | Tissue | RT-PC |
| Xu etal | CIRCNFIC ↓ | 2020 | China | 150 | 2.81 | 0.26-29.83 | 2.0FC | Tissue | RT-PC |
| Liu etal | CircRNA_103809 ↓ | 2020 | China | 65 | 1.06 | 0.3-3.85 | Median | Tissue | RT-PC |
| Li etal | Circ-VRK1 ↑ | 2019 | China | 350 | 2.08 | 1.07-4.03 | Median | Tissue | RT-PC |
| wang etal | CircZNF609 ↑ | 2018 | China | 143 | 1.16 | 0.44-3.03 | Median | Tissue | RT-PC |
| Zhang etal | circ-LARP4 ↓ | 2019 | China | 283 | 1.94 | 1.85-4.45 | Median | Tissue | RT-PC |
| Mao etal | circ000554 ↓ | 2020 | China | 138 | 1.63 | 0.8-3.32 | Mean | Tissue | RT-PC |
| Zang etal | circ_0000520 ↑ | 2020 | China | 60 | 1.5 | 0.18-12.5 | Median | Tissue | RT-PC |
| Chen etal | CirCHIPK3 ↑ | 2020 | China | 50 | 1.69 | 0.14-20.22 | Median | Tissue | RT-PC |
| Lin etal | TV-circRGPD6 ↓ | 2020 | China | 217 | 2.45 | 1.5-4.01 | 2.0FC | Tissue | RT-PC |
| Bian etal | Circular PVT1 ↑ | 2020 | China | 99 | 1.6 | 0.64-4 | Median | Tissue | RT-PC |
| Cai etal | circ0000515 ↑ | 2020 | China | 340 | 1.39 | 0.69-2.82 | Mean | Tissue | RT-PC |
| Li etal | Hsa_circ_002178 ↑ | 2020 | China | 83 | 1.34 | 0.32-5.62 | Median | Tissue | RT-PC |

BC breast cancer; ↑ or ↓ up-regulated or down-regulated with poor prognosis; OS overall survival; HR hazard ratio; NR not report; FC fold change ;RT-PCR reverse transcription PCR.

**Table 2:**
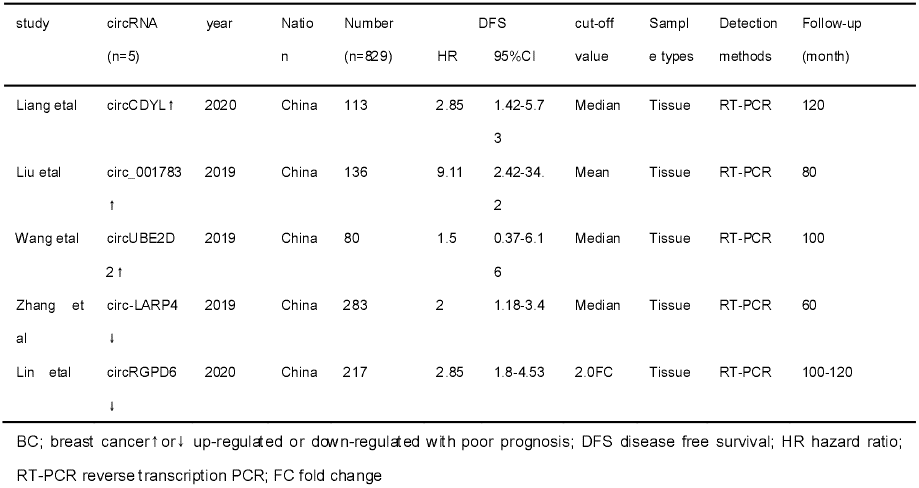
Characteristics of studies and different circRNAs expression related to DFS in BC patients.

**Table 3:**
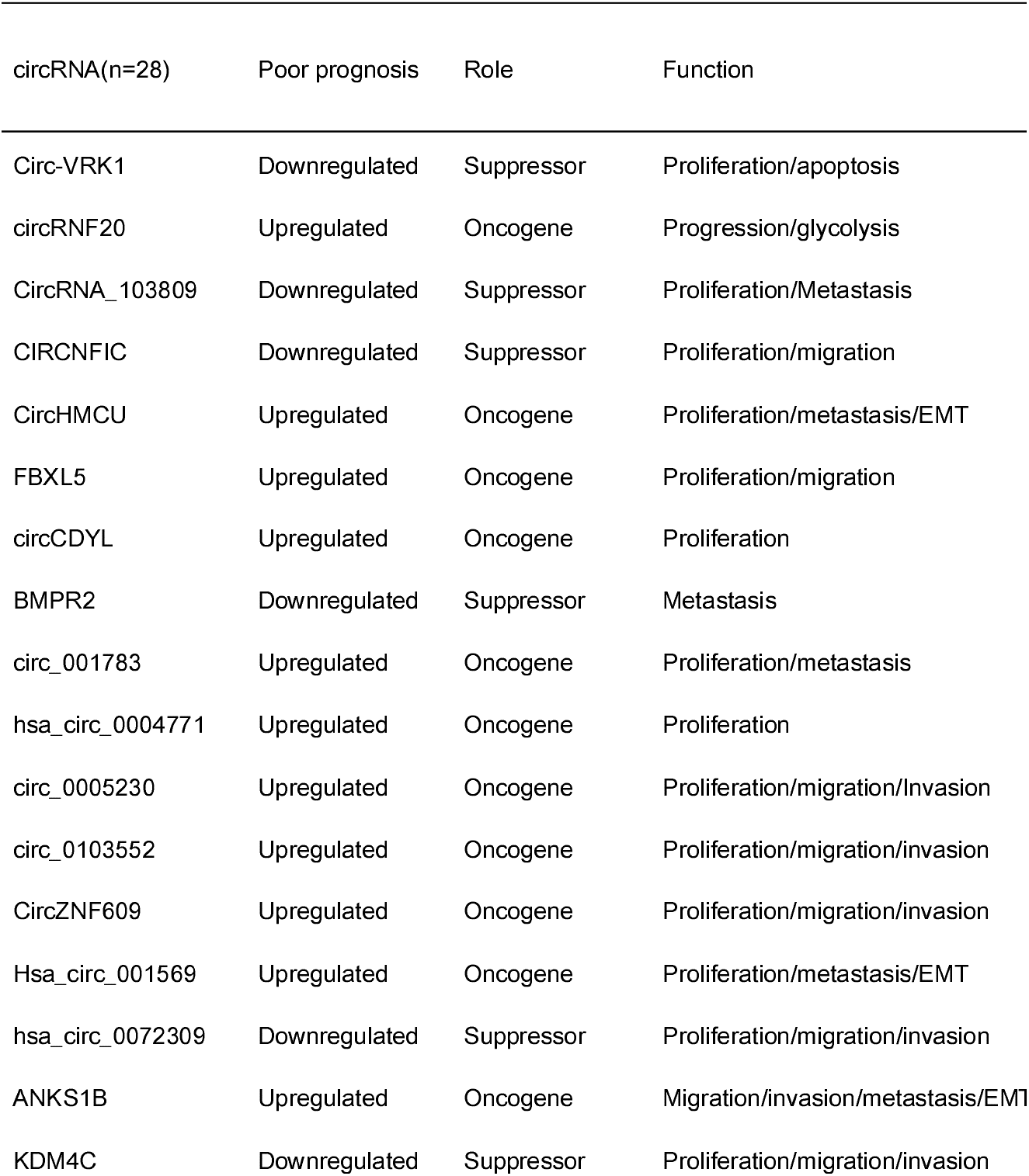
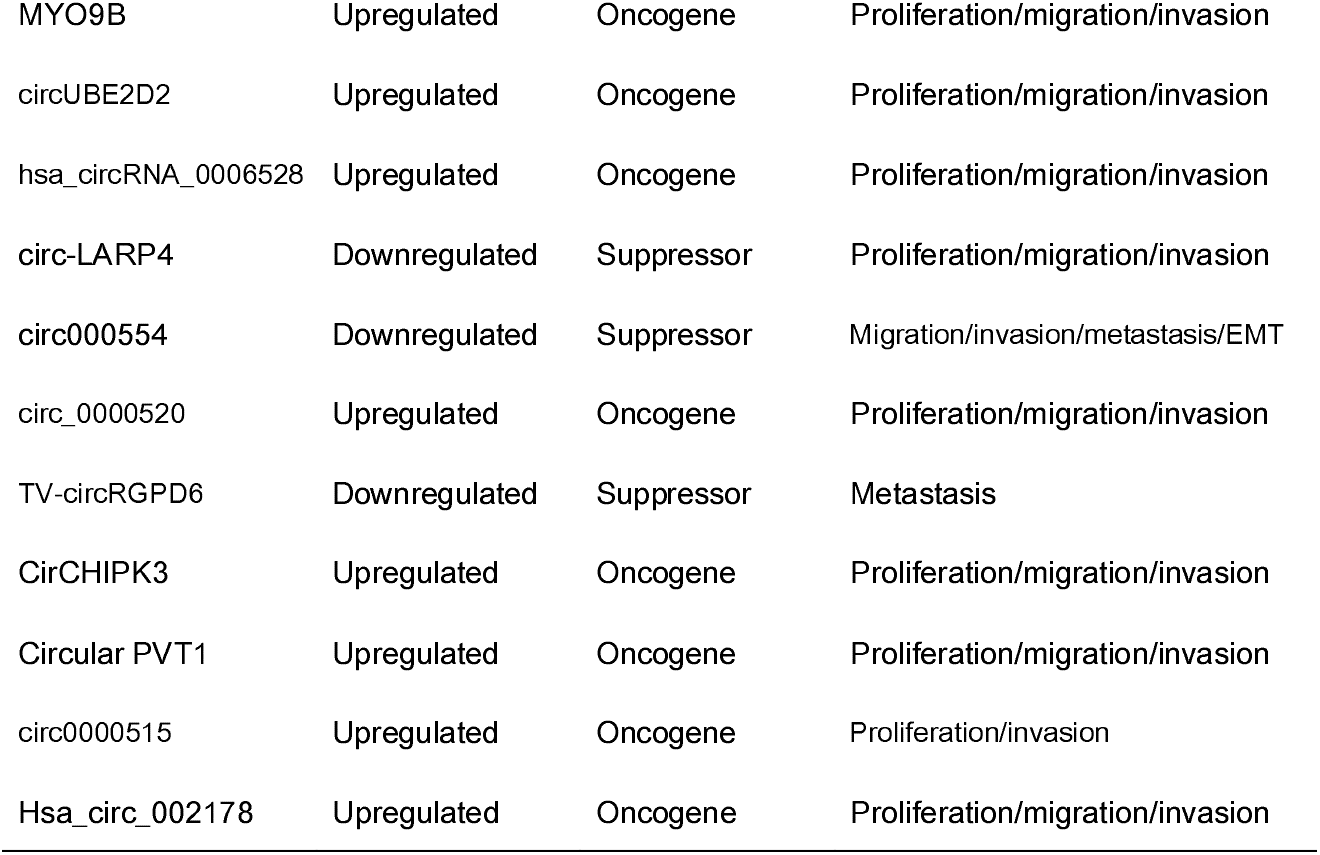
circRNAs and target miRs in breast cancer.

### 3.3. Meta-Analysis Findings

Random-effects and fixed-effects models were applied to evaluate the pooled hazard ratio (HR) and its corresponding 95% confidence interval (CI) of OS or DFS based on the heterogeneity level. The pooled HR value (95% CI) of OS which correlated with the expression of circRNA was 1.68 (1.44-1.97) with moderate heterogeneity (I^2^=47.9%,P=0.003) and statistically significant (P < 0. 00001) (Figure 2).Therefore,the random effects models was used. And the pooled HR value (95% CI) of DFS related to different circRNAs expressions was 2.63 (1.95-3.53) in BC with low heterogeneity (I^2^=22.9%, P=0.268) and statistically significant (P < 0 .00001) (Figure 3). As is showed in figure 4, 28 included studies were divided into six subgroups by the cut-off value.We found that 2.0FC subgroup(I^2^=87.1%) was the main source of heterogeneity.Furthermore, funnel plots of included studies relevant to circRNAs, OS, and DFS in BC patients were presented in Figures 5, and 6, respectively. These figures are approximately symmetrical, and we can think that there is no obvious publication bias.

**Figure 2:**
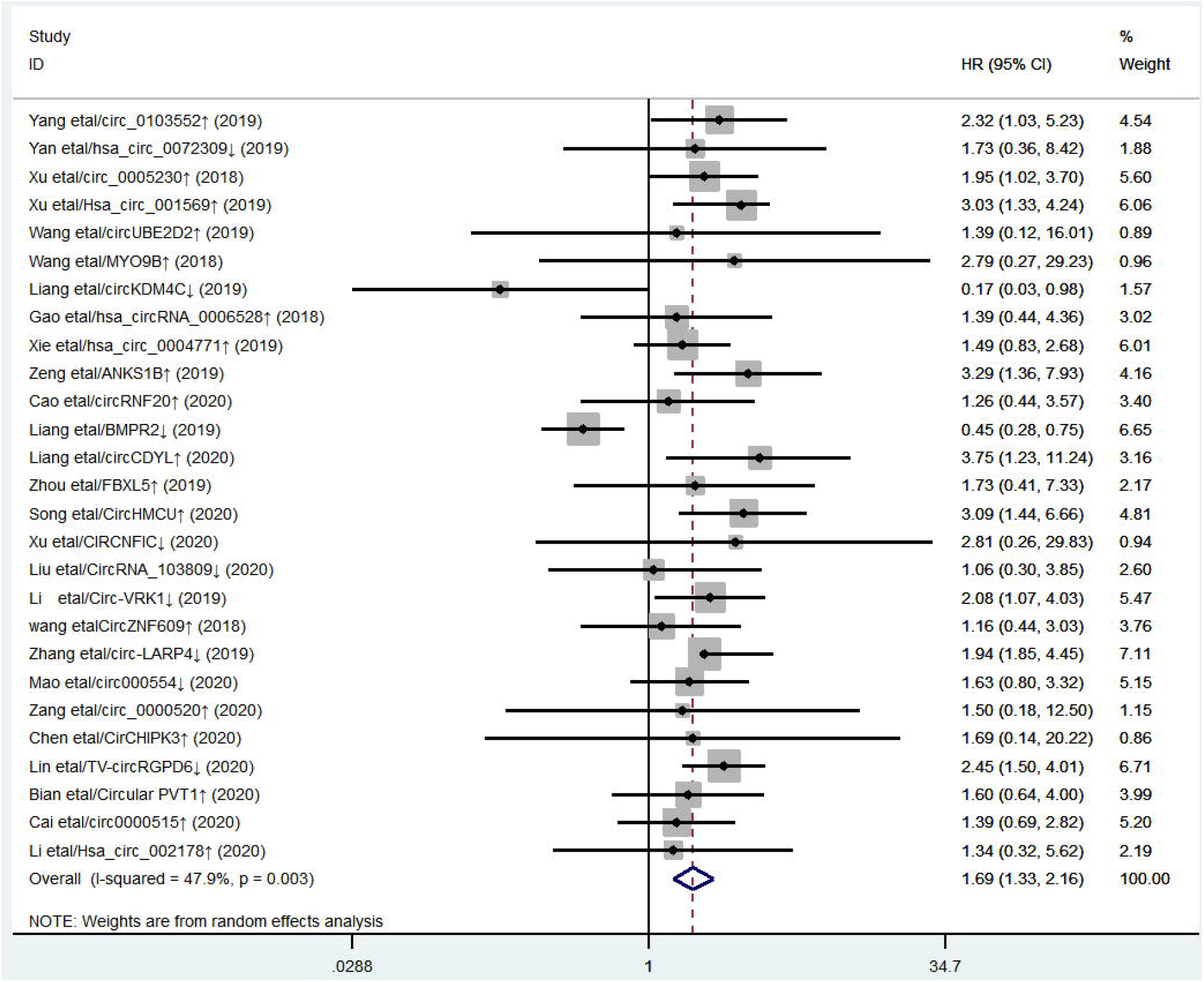
Forest plot of OS associated with expression level of different circRNAs in BC patients was presented.

**Figure 3:**
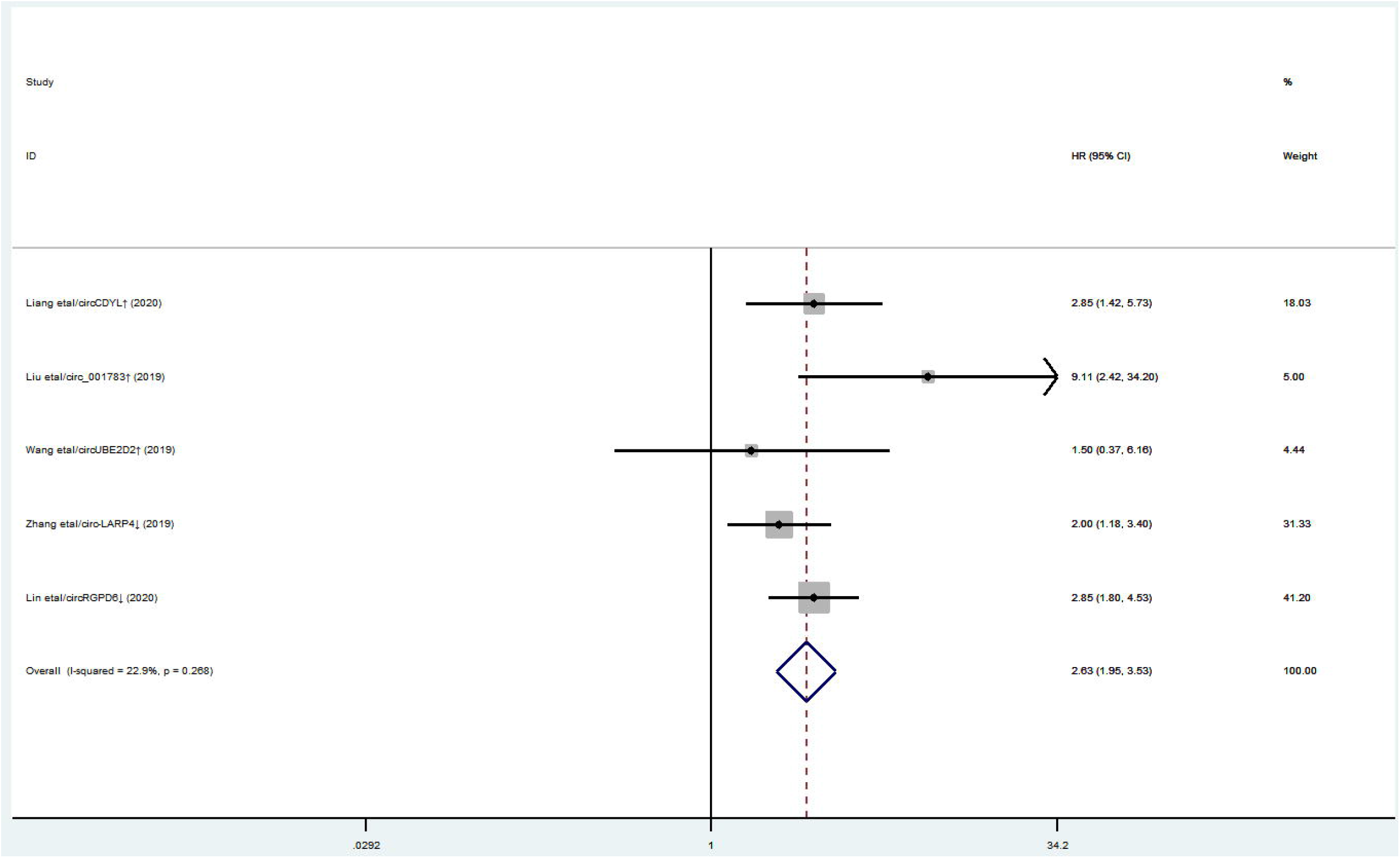
Forest plot of DFS associated with expression level of different circRNAs in BC patients was presented.

**Figure 4:**
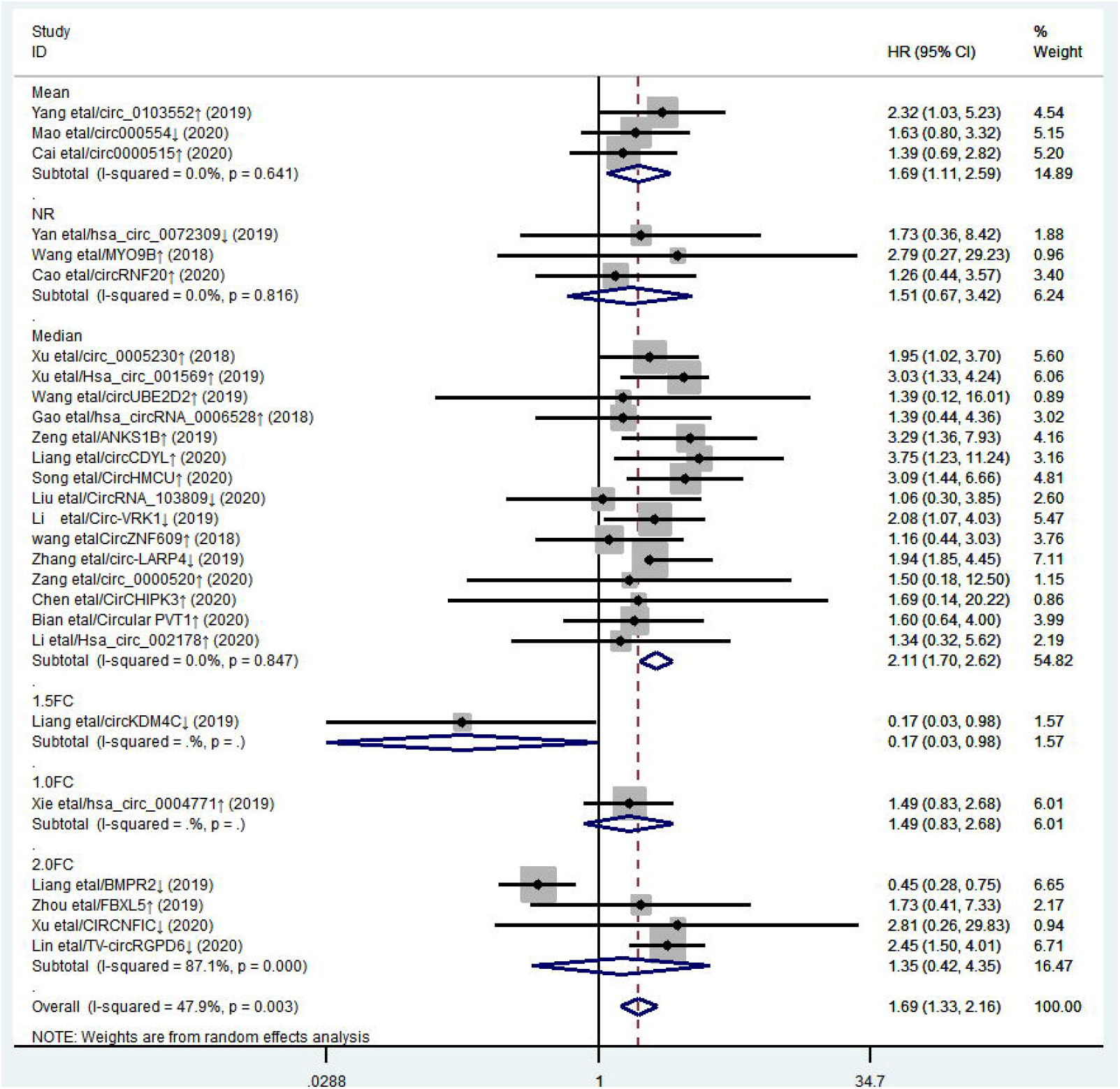
Forest plot of OS associated with expression level of different circRNAs in BC patients by subgroups

**Figure 5:**
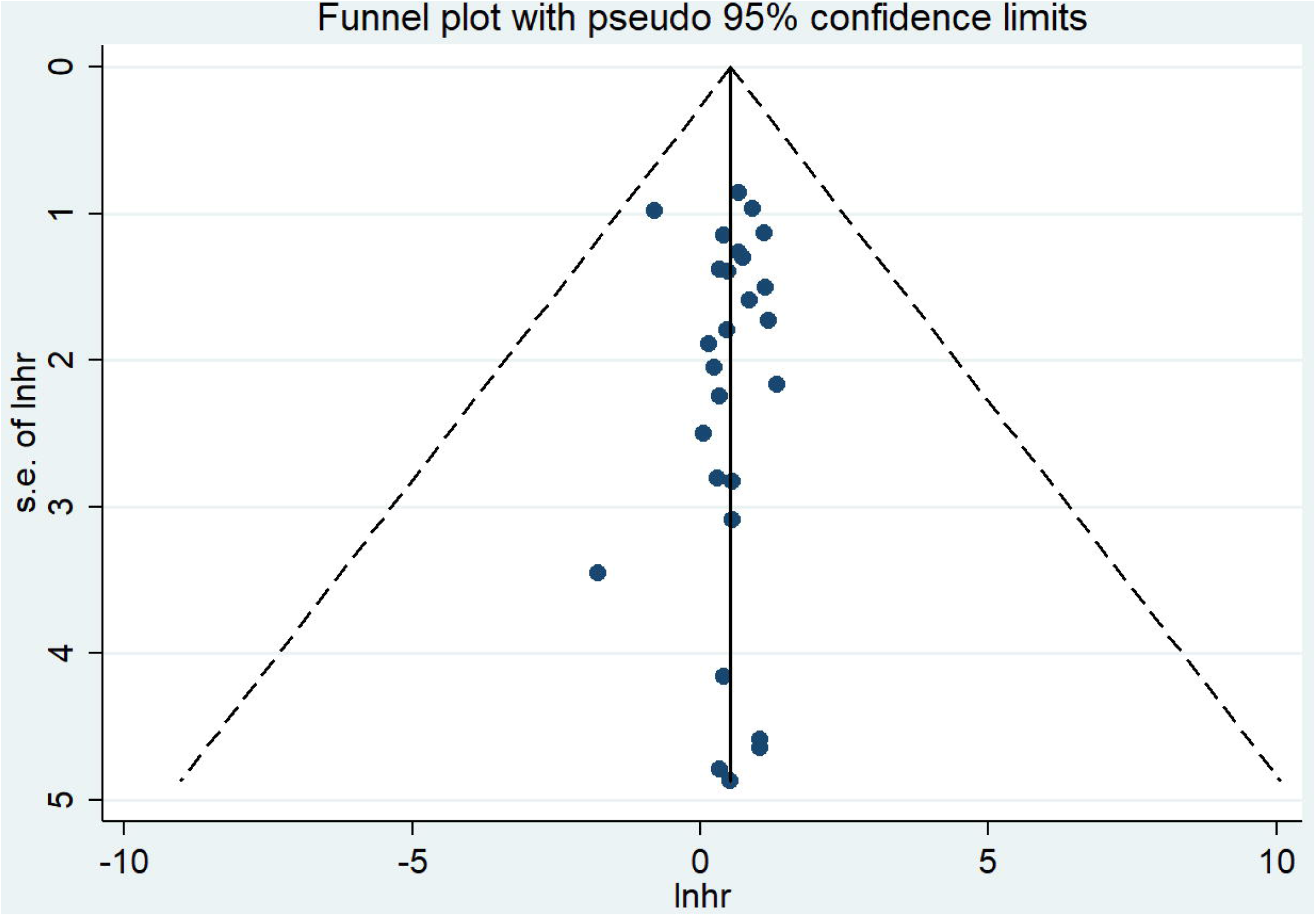
Funnel plots of studies associated with OS in this meta-analysis.

**Figure 6:**
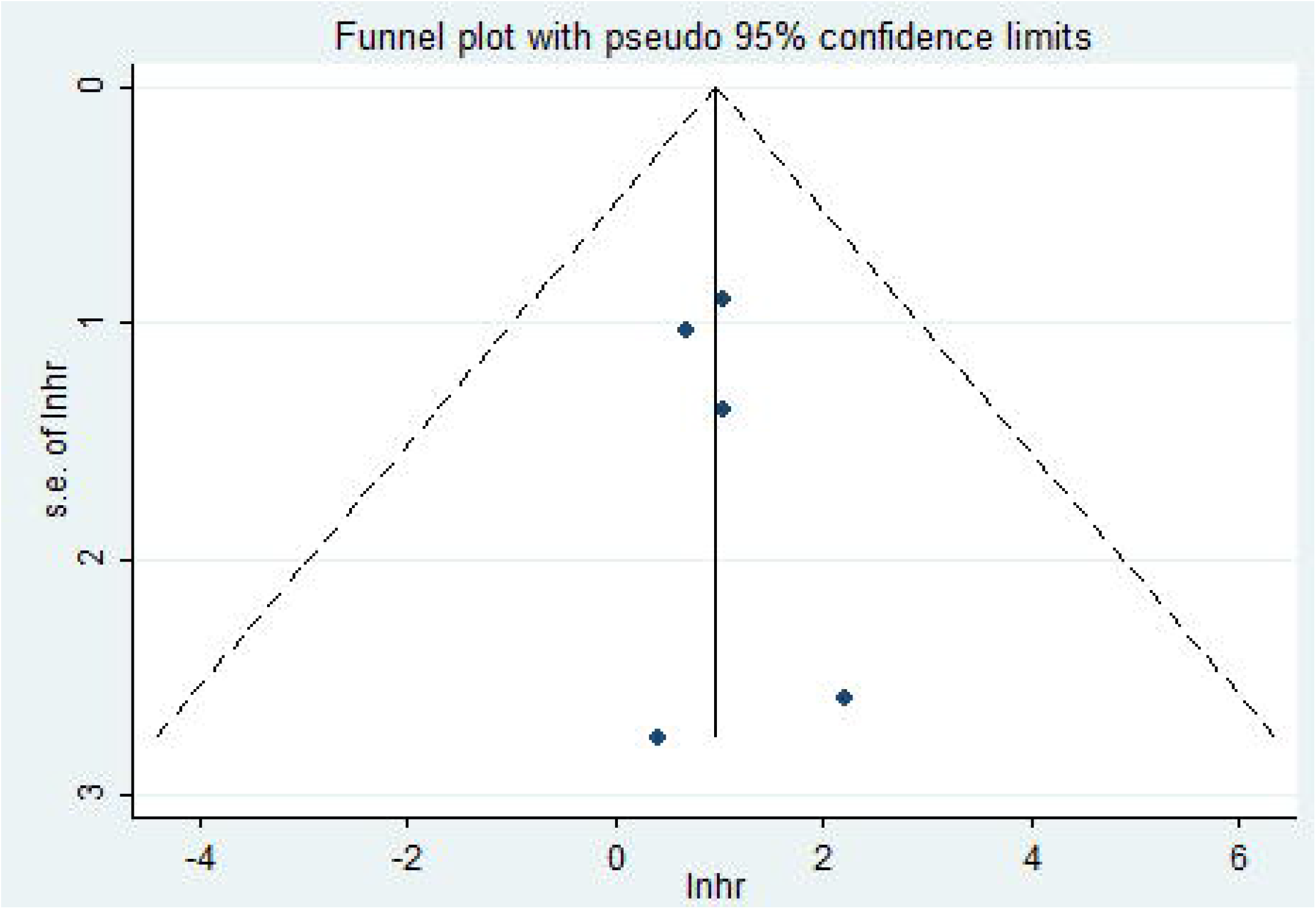
Funnel plots of studies associated with DFS in this meta-analysis.

## 4. Discussion

Breast cancer remians a huge threat to human health, although new anticancer drugs are ongoing emergence on account of chemotherapy resistance and metastasis inducing poor prognosis.In the last decade, more and more studies focused on the clinical roles of circRNAs and many reports indicated that circRNAs can be a molecular biomarker in BC patients for predicting prognosis. However, few relevant meta-analysis on the prognostic value of circRNAs expression existed in BC.

The purpose of this study was to elucidate the relationship between circRNAs expression and prognosis in breast cancer patients. Through big data meta-analysis, we provided evidence to demonstrate the prognostic value of abnormally expressed circRNAs in BC patients. The results of this meta-analysis showed that the pooled HR values (95%CI) of OS and DFS associated with different circRNAs expression in BC patients were 1.68 (1.44-1.97) and 2.63 (1.95-3.53), respectively, which suggested that the abnormally expressed circRNAs could be used as a cancer biomarker in BC patients. By detecting specific circRNAs expression levels in tissues or other body fluids, we can not only make appropriate clinical decisions based on different prognoses, but also monitor the treatment outcomes of BC patients receiving different treatments. The main role of circRNAs at the molecular level is to act as microRNA (miRNA) sponges and isolate miRNAs, thereby terminating the regulation of miRNA target genes [39, 40].

There is increasing evidence [41-46]that circRNAs play a key role in various cellular activities and are involved in various human diseases, including the proliferation, migration, invasion and cell cycle-related activities of cancer. The upregulation of circRNAs promoted the migration, invasion, EMT, proliferation and drug resistance of tumor cells, and inhibited the apoptosis of different target genes, which was related to poor prognosis. For example, Zang et al.[32]reported that circ_0000520 gene knockdown could inhibit the growth, migration and invasion of cells in vitro and in vivo by directly regulating Mir-1296 /SP1 pathway. The results showed that circ_0000520 was up-regulated in breast tumors and cell lines, and its high expression was associated with low overall survival rate. The down-regulated circRNAs with overexpression can inhibit the migration, invasion and proliferation of tumor cells. Yan Ling et al.[25]found that circ_0072309 with overexpression could inhibit the proliferation, metastasis and invasion of breast cancer cells by inhibiting the Mir-492 pathway, and the results showed that overall survival rate of circ_0072309 with low expression was high campared to circ_0072309 with overexpression.

Therefore, circRNAs can be used as a molecular biomarker to predict the prognosis of BC patients. Several studies have shown that circRNA can be used as CeRNAS to regulate the proliferation and progression of BC through micro RNA sponges. There is increasing evidence that micrornas regulate gene expression during tumorigenesis. further studies have been found that some circRNAs, as microRNA sponges, are involved in the regulation of tumor proliferation, metastasis and invasion. For example,Bian et al. [34] found that Circular PVT1 was significantly overexpressed in breast cancer tissues and cell lines, and it plays a role of competitive endogenous RNA (CE RNA) by targeting Mir-204-5P. Circular PVT1 knockdown can inhibit the proliferation, invasion and metastasis of breast cancer cells. This study explored how the ceRNA mechanism of circRNA functioned in the oncogenesis and progression of breast cancer,providing a new perspective for basic research.In general, predicting patient outcomes and exploring the mechanisms of circRNA play a key role in clinical decision-making and the development of new targeted gene therapies.

we summarized the research on the mechanism of circRNA. We found that 28 studies showed specific targets for circRNAs, 9 circRNAs were down-regulated, and their overexpression could inhibit the proliferation, infiltration or metastasis of breast cancer cells. Nineteen circrnas were up-regulated, and their overexpression could promote proliferation, invasion or metastasis of breast cancer cells. These studies have shown that the potential relationship between circRNAs and microRNA plays a key role in the pathogenesis of tumors and promotes the research and development of oncogene therapy. Many studies focusing on the same circRNAs have revealed different targets, and the potential correlation between circRNAs and microRNA remains unclear. In the future, we should pay attention to the interaction between circRNAs and microRNA or other types of RNA, and achieve targeted therapy by intervening multiple types of RNA simultaneously.

However, there are some limitations in this study. First, most of the patients are from East Asia, especially China, which makes our conclusion only suitable for Chinese patients. Second, after the exclusion of the 2.0FC subgroup, meta analysis revealed a significant decrease in heterogeneity, which suggested that different cut-off values and detection methods for circRNAs expression in different studies may lead to heterogeneity among studies. Third, language bias is also a limitation, as we only register English papers in meta-analysis. Fourth, most authors are generally more likely to report positive results, so the calculated aggregate effect value may overestimate the predictive significance of circRNA in the prognosis of breast cancer patients. Consensus has been reached on publication bias. Fifth, we calculated HR estimates from the Kaplan-Meier survival curve, because some studies could not directly extract HR and 95%CI. Sixth, some confounding factors in studies may lead to high heterogeneity if there is no adjusted HR value.

In conclusion, this meta-analysis supports the fact that specific circRNAs is significantly associated with the prognosis of breast cancer patients and may serve as a new marker for predicting the prognosis of breast cancer patients. In addition, circRNAs may contribute significantly to future circRNAs targeted therapy and clinical decision-making for breast cancer.

## Data Availability

literature search was performed in the PubMed, Scopus, EMBASE, and the Cochrane Library databases

